# The role of alternative splicing in CEP290-related disease pathogenesis

**DOI:** 10.1101/2022.03.03.22271834

**Authors:** Rowan D. Taylor, James A. Poulter, Joseph Cockburn, John E. Ladbury, Michelle Peckham, Colin A Johnson

## Abstract

Primary ciliopathies are a group of inherited developmental disorders resulting from defects in the primary cilium. Mutations in *CEP290* (Centrosomal protein of 290kDa) are the most frequent cause of recessive ciliopathies (incidence up to 1:15,000). Pathogenic variants span the full length of this large (93.2kb) 54 exon gene, causing phenotypes ranging from isolated inherited retinal dystrophies (IRDs; Leber Congenital Amaurosis, LCA) to a pleiotropic range of severe syndromic multi-organ ciliopathies affecting retina, kidney and brain. Most pathogenic *CEP290* variants are predicted null (37% nonsense, 42% frameshift), but there is no clear genotype-phenotype association. Almost half (26/53) of the coding exons in *CEP290* are in-phase “skiptic” (or skippable) exons. Variants located in skiptic exons could be removed from *CEP290* transcripts by skipping the exon, and nonsense-associated altered splicing (NAS) has been proposed as a mechanism that attenuates the pathogenicity of nonsense or frameshift *CEP290* variants. Here, we have used *in silico* bioinformatic techniques to study the propensity of *CEP290* skiptic exons for NAS. We then used CRISPR-Cas9 technology to model *CEP290* frameshift mutations in induced pluripotent stem cells (iPSCs) and analysed their effects on splicing and ciliogenesis. We identified exon 36, a hotspot for LCA mutations, as a strong candidate for NAS that we confirmed in mutant iPSCs that exhibited sequence-specific exon skipping. Exon 36 skipping did not affect ciliogenesis, in contrast to a larger frameshift mutant that significantly decreased cilia size and incidence in iPSCs. We suggest that sequence-specific NAS provides the molecular basis of genetic pleiotropy for *CEP290*-related disorders.

## Introduction

Primary cilia are microtubule-based organelles that protrude from the cell surface of most mammalian cells during G0 and G1 of the cell cycle, acting as important signalling hubs. Cilia participate in diverse roles in chemo-, mechano- and photosensation allowing us to hear, see, excrete, and reproduce. They have essential roles in vertebrate development and growth factor signalling pathways (including Shh, PDGF, Wnt), and an ever-expanding group of developmental disorders known as ciliopathies are caused by defects in ciliary structure or function. As a group, ciliopathies, including autosomal dominant polycystic kidney disease, are common Mendelian inherited conditions that have an estimated incidence of 1 in 2000 (1). Pathogenic variants in at least 190 genes cause ciliopathies, encompassing vast genetic and phenotypic heterogeneity, with over 30 separate described clinical entities (2). Frequent clinical features include vision loss, kidney failure due to renal cystic dysplasia, infertility, obesity, skeletal malformations, and structural brain abnormalities.

CEP290 (Centrosomal protein of 290kDa) is a 93.2kb, 54 exon gene that encodes a 290kDa protein of 2479 amino acids (aa). It is a key regulator of ciliary content and cilia formation (3). Mutations in *CEP290* (MIM610142) cause a wide range of autosomal recessive ciliopathies, ranging from the least severe isolated retinal diseases Retinitis Pigmentosa (RP) (MIM268000) and Leber Congenital Amaurosis (LCA) (MIM611755), through to the embryonic lethal syndromic disease Meckel-Gruber Syndrome (MKS) (MIM611134). Other multi-organ diseases caused by disruption to *CEP290* include Senior Loken syndrome (SLS) (MIM610189), Joubert syndrome and related disorders (JBTS and JSRD) (MIM610188), and Bardet Biedl syndrome (BBS) (MIM615991).

*CEP290* has over 200 associated pathogenic variants spread throughout the full length of the gene. There are also 136 affected probands in the 100,000 Genomes Project datasets with biallelic likely pathogenic CEP290 variants and ciliopathy phenotypes (4, 5). However, there is no clear genotype-phenotype association between severe syndromic and isolated retinal disease-causing genotypes. The majority of pathogenic *CEP290* mutations are predicted null alleles (37% nonsense, 42% frame-shifting) that introduce premature stop codons, predicted to cause complete loss of full-length protein through nonsense-mediated decay (NMD) (6). However, for some of these variants, even in homozygous form, affected individuals display non-syndromic retinal disease, rather than the expected multi-organ defects (as is the case for JSRD, BBS and MKS) predicted to be caused by null alleles.

One hypothesis for genetic pleiotropy of *CEP290* disease has been the existence of modifier alleles, which have been shown to enhance or dampen disease phenotype in some cases (7, 8). However, this does not fully explain the broad pleiotropy exhibited by *CEP290* mutations. An alternative theory posits that some exonic variants in *CEP290* may trigger exon skipping during splicing, in a tissue-specific manner (9-12). In these instances, in-frame exons (i.e., those with n/3 nucleotides, containing a nonsense or frameshift variant, can be spliced out from the gene transcript via basal exon skipping or nonsense-associated altered splicing (BES/NAS). This would restore the reading frame and produce a truncated protein that retains some or all of the functionality of the wild-type protein (13). The amount of truncated protein depends on the efficiency of BES/NAS, which is presumed to be exon and cell-type dependent.

This has been suggested for *CEP290*, but also other ciliopathy disease genes, such as *CC2D2A*, and unrelated genes such as *BRCA2* (9, 14, 15). A key example of splicing-mediated pleiotropy in *CEP290-*related disease is the deep intronic variant c.2991+1665A>G, which introduces a cryptic exon containing a premature stop codon and causes an LCA phenotype. A residual amount of *CEP290* transcript is still spliced correctly (without the cryptic exon). This accounts for the lack of disease pathogenicity in other organs, as the amounts of residual full length CEP290 protein can be as high as 50% in patient fibroblasts. However, the retina is more sensitive to loss of CEP290 protein than in other organs for which lower levels are sufficient to maintain cilia function (9, 16). The large number of in-frame exons within *CEP290* (n=26/55) mean that many of them could theoretically be skipped, whilst maintaining the frame of the transcript to produce near-functional albeit truncated protein.

If *CEP290* exonic variant candidates for induced exon skipping can be identified, this opens the possibility of personalised antisense oligonucleotide (AON) treatment to treat patients with these disease-causing variants. Experimental interventions have shown promising results in cell and animal ciliopathy models by AONs to attenuate disease phenotype through forced skipping of skiptic variant-containing exon(s) (10, 17, 18). However, tissue specificities have not been demonstrated in these studies raising the question of whether NAS is both sequence- and tissue-specific. This has important implications for the efficacy and delivery of any potential splice switching AONs.

In this study, we analysed the human exome for genes that had high prevalence of in-frame exons and enriched protein domains, GO terms, and disease types associated with these genes. We selected *CEP290* as a candidate gene for further analysis because it had a high prevalence of in-frame exons with enrichment of coiled-coil domains and mutations cause broad pleiotropy that are suitable for assessment of exon skipping. We then assessed the natural differential exon usage between key *CEP290-*affected organs by RNASeq analysis of public datasets. Reported *CEP290* pathogenic nonsense variants were collated, with patient genotypes and associated diseases, and *in silico* structural predictions were made, in order to identify hotspot regions for different disease types. *In silico* splicing predictions for these variants identified skiptic exon 36 (ENSE0001666646) as a hotspot for LCA-causing mutations that are candidates for nonsense-associated altered splicing (NAS). We modelled biallelic exon 36 variants using CRISPR-Cas9 gene editing in human induced pluripotent stem cells (iPSCs), demonstrating variable effects on exon skipping, transcript usage, protein expression and ciliogenesis.

## Results

### In-frame exon representation in the human exome

We generated a library of human exons (n=353,184 exons with a unique Ensembl ID and InterPro annotation, for a total of n=19,135 genes with GO term annotation) linked to their flanking intron phases (exon-phase). Over 10% of exons are of the (0, 0) symmetric exon type (n=35,628), and these are: (1) significantly enriched for the InterPro term “coiled-coil domain” (p=5.01×10^−37^ hypergeometric test; expect 1032.5, actual 1429), suggesting that exons encoding coiled-coil domains are often symmetric (Figure 1).

**Figure 1.**
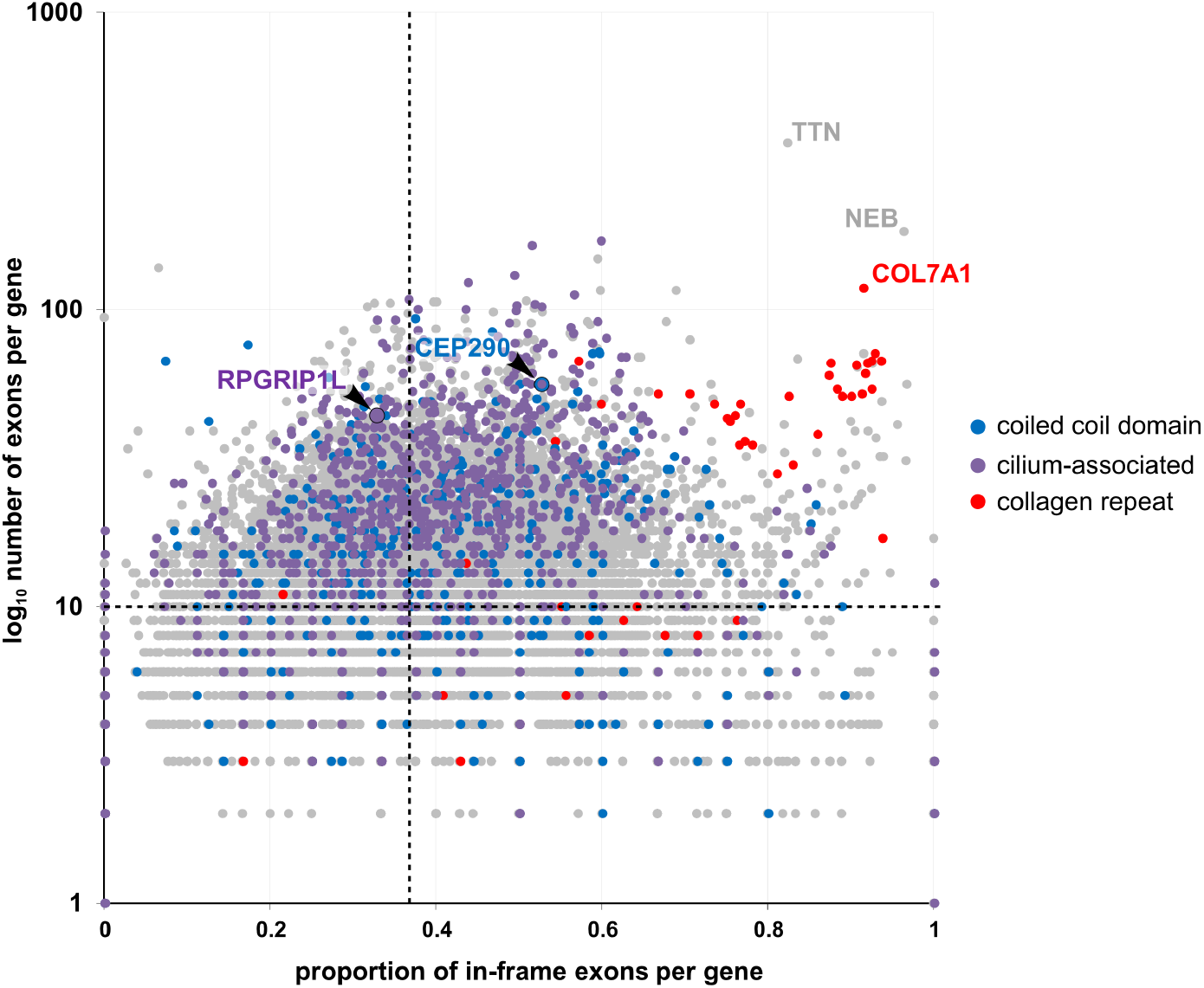
In-frame exon representation in human exome. Graphical representation of proportion of in-frame exons in all human genes against the gene exon number (log10 scale). *Purple dots* - positive for GO terms “cilium” or “basal body”, *red dots* - positive for InterPro term “collagen triple helix OR repeat”, *blue dots* – positive for InterPro term “coiled-coil domain”. Dashed lines represent mean proportion of in-frame exons (0.362) and mean number of exons per gene (10.705).

The mean proportion of exons that are in-phase = 0.362, and genes with a greater than average proportion are enriched for GO terms (2) “retinal eye development” (p=1.56×10^−3^ hypergeometric test; expect 75.2, actual 87) and (3) “cilium OR basal body” (p=1.05×10^−3^; expect 112.2, actual 139). The intersection of the three search terms only identifies the ciliary proteins RPGRIP1L and CEP290. 51% of the exons in the canonical *CEP290* transcript (ENST00000552810.6) are in-frame and *CEP290* mutations cause hugely pleiotropic ciliopathy phenotypes. These analyses provide our initial rationale for further studies of *CEP290*.

### Naturally occurring alternative splicing of CEP290 mRNA

To determine tissue-specific splicing differences in the affected tissue types in *CEP290-*related disease, native differential exon usage was first analysed. RNAseq data was gathered from NCBI’s Short Read Archive from unaffected adult human cerebellum, brain, kidney, retina, thyroid, and skin samples. Exon numbers throughout refer to the canonical transcript ENST00000552810.6, unless otherwise stated.

The relative exon usage between samples was compared. Significantly differentially expressed exons were situated throughout *CEP290* (Figure 2A and B). Two of the 21 significantly differentially expressed exons are untranslated first exons of the canonical transcript (which is shared with one other protein coding transcript) and the alternative transcript ENST00000672647.1 (untranslated transcript). Interestingly, for exon 1a, which is only present in ENST00000672647.1, it has significantly higher expression in the retina than in the kidney.

**Figure 2.**
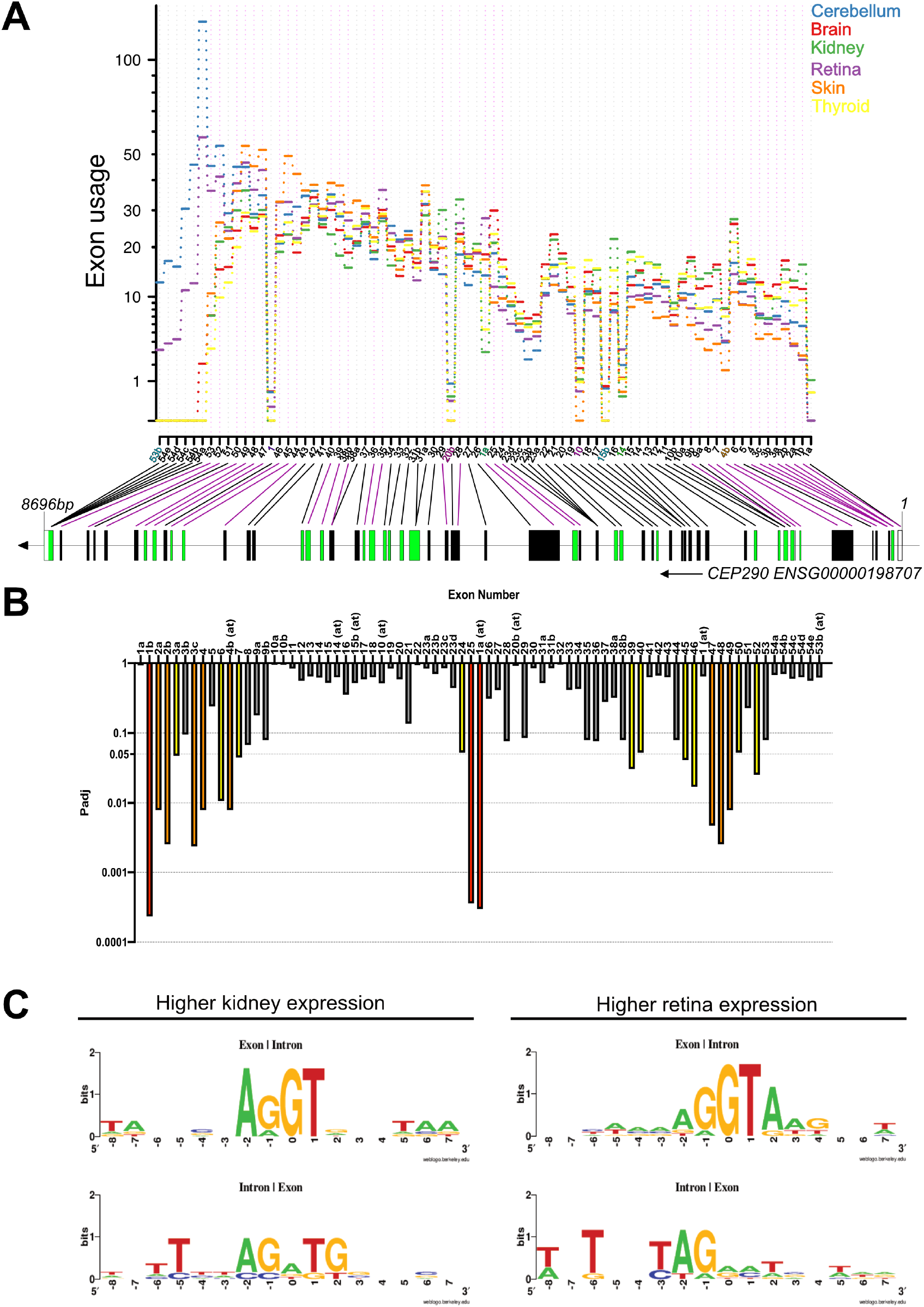
*CEP290* exon usage analysis. A. *CEP290* relative exon usage values in disease-relevant organs. Skin – control organ. Data analysed, graph produced and adapted from DEXSeq(20). Values recorded for counting bins on the x axis representing whole exons or regions of exons as indicated in the gene diagram below. Coloured counting bins represent exons from alternative *CEP290* transcripts. Green boxes – symmetric exons. B. Adjusted p values presented graphically to show significant differential expression of exons as calculated by DEXSeq (Benjamini-Hochberg correction, False Discovery Rate = 0.1). Highlighted bars: *yellow* – 0.05>p_adj_>0.01, *orange* 0.01>p_adj_>0.001, *red* – 0.001>p_adj_. C. Splice consensus site alignment of exons at their 5’ and 3’ intron boundaries with significantly higher kidney expression vs. higher retinal expression.

Of the 19 significantly differentially expressed coding exons, 12 were in-frame exons (2, 3, 6, 4b, 7, 25, 39, 40, 45, 46, 47 and 48) and 5 were not in-frame (4, 24, 49, 50 and 52). However, exons 24, 49 and 50 are all fully or partially missing from the 1469aa protein-coding alternative transcript ENST00000547691.8. Exon 4b is the 2^nd^ half of an exon included in transcript ENST00000552770.3 (a 154aa protein-coding transcript), the first half of this exon shares sequence with exon 6 in the canonical transcript, which is also significantly differentially expressed. Exons 4b and 6 both have significantly greater expression in the brain and kidneys than the skin (unaffected control organ).

CEP290 is structurally unresolved, but the locations of exons can be mapped to predicted structural/functional domains. Exons 4-7 are situated in the 1^st^ putative coiled-coil domain, within the N-terminal homo/heterodimerisation domain and are all within the predicted cellular membrane-binding domain (4, 19). Exons 24 and 25 are within the 4^th^ putative coiled-coil domain and the structural maintenance of chromosomes (SMC) homology domain, while exons 47-54 are in the 13^th^ and final predicted coiled-coil domain and the C-terminal homo/heterodimerisation region (19).

To assess whether splicing differences in CEP290 were apparent between tissue types, significantly differentially expressed exons were grouped into those that had higher retinal expression and those that had higher kidney expression compared with other tissue types. The exon-intron and intron-exon boundaries were aligned for each group and the consensus sites compared. Clear differences in consensus sites between the two groups were observed, particularly at the intron-exon boundaries (Figure 2C). This is indicative of differences in alternative splicing between the two tissue types.

### NAS candidate variant identification

Disease-causing variants in *CEP290* are commonly deleterious (frameshift or nonsense) (Figure 3A). The distribution of exonic nonsense variants by disease type, predicted structural domains, and binding sites of potential interacting proteins are illustrated in Figure 3B. 53% of the *CEP290* nonsense variants listed on ClinVar are in exons that are in-frame and/or missing from naturally occurring alternative transcripts. A hotspot of nonsense variants within skiptic exon 36 causes non-syndromic retinal disease (Leber Congenital Amaurosis – LCA) and is therefore likely to be a strong candidate exon for NAS. In the encoded protein, our *in silico* structural analysis of CEP290 found that over 56% of the protein contains 29 predicted coiled coil domains separated by regions of unknown structure. Exon 36 is predicted to be in a link region between two coiled coil domains in the encoded protein.

**Figure 3.**
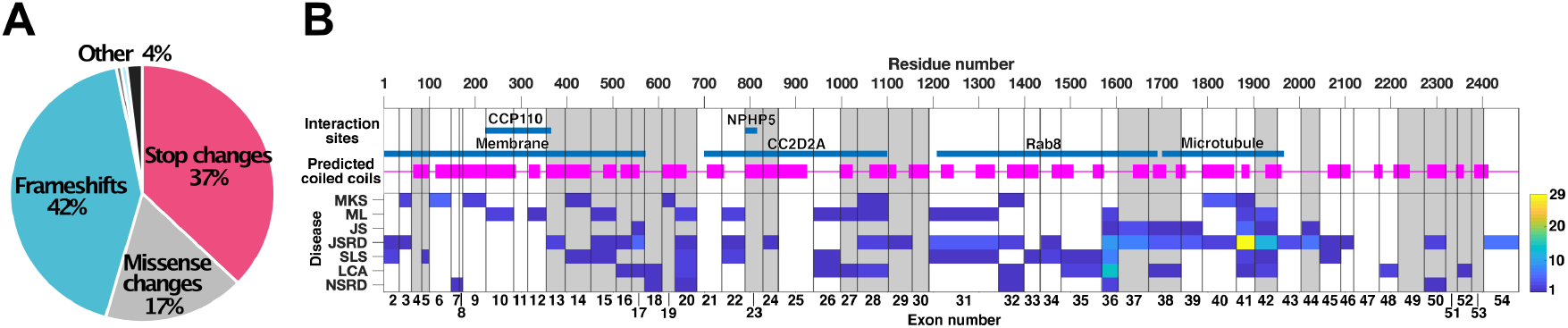
Prevalence and distribution of *CEP290* disease variants. A. Mutation types of unique pathogenic variants in *CEP290*. Data source: LOVD (https://databases.lovd.nl/shared/genes/CEP290/graphs, accessed September 2020). B. Distribution of nonsense variants in *CEP290*, predicted interactant binding sites, and putative coiled coils. Coding exons 2 to 54 are indicated by vertical lines, symmetric exons are white. Teal lines indicate putative binding sites or interaction regions. Predicted coiled coil domains highlighted by magenta blocks. Heat-map of position and number of known ClinVar pathogenic variants, mapped by ciliopathy disease phenotype (MKS, Meckel-Gruber syndrome; ML, MKS-like; JS, Joubert syndrome; JSRD, JS & related disorders; SLS, Senior-Loken syndrome; LCA, Leber congenital amaurosis; NSRD, non-syndromic retinal dystrophy, either severe early childhood onset retinal dystrophy (SECORD) or early onset retinitis pigmentosa (EORP). Hot-spots (blue-yellow) of pathogenic nonsense variants occur in symmetric exon 6 (causing MKS), exon 36 (LCA) and exons 41 (JSRD) highlighted in red. Other symmetric exons of interest are 9 & 40.

We aimed to collate all known pathogenic nonsense variants in *CEP290* and to determine their potential for exon skipping that could ameliorate phenotype. Patient genotypes and their associated phenotypes were data-mined from the literature for all NCBI ClinVar nonsense *CEP290* variants (Supplementary Table 1) and splicing predictions were carried out (Supplementary Table 2).

Many *CEP290* disease patients are compound heterozygous for pathogenic *CEP290* variants. As a consensus, when one allele contains a stop variant and the other contains a “less severe” variant such as a splicing or missense variant, this results in retina-specific disease rather than syndromic disease, consistent with the central role of CEP290 in photoreceptors (21). However, there are also instances with biallelic pathogenic nonsense variants (either homozygous or compound heterozygous) causing retina-specific disease rather than syndromic disease.

Our results show a high incidence of nonsense variants that are predicted to significantly alter splicing in LCA hotspot, skiptic exon 36. The prevalent disease-causing variant c.4723A>T, p.Lys1575Ter (7), causes LCA in homozygous form. However, in compound heterozygous form with other nonsense variants causes syndromic diseases SLS or JSRD, with some exceptions. HOT-SKIP and Human Splicing Finder (HSF) predict significant alteration to the exonic splice enhancer / exonic splice silencer (ESE/ESS) ratio resulting from c.4723A>T. Homozygous c.4771C>T, p.Gln1591Ter also causes LCA, and both HOT-SKIP and HSF predict significant alteration to ESE/ESS ratio. Furthermore, c.4801C>T, p.Gln1601Ter which has been found in an LCA patient, compound heterozygous for c.4801C>T and the common deep intronic variant c.2991+1665A>G, is also predicted to significantly alter the ESE/ESS ratio by HOT-SKIP and HSF. Interestingly, the other homozygous nonsense mutations in exon 36 (c.4732G>T, p.Glu1578Ter and c.4811G>A, p.Trp1604Ter) both cause syndromic Joubert syndrome (JBTS) and are not predicted to significantly alter splicing.

### Mutations in LCA hotspot CEP290 exon 36 show sequence-specific altered splicing in cell models

We have observed that exon 36 of *CEP290* is a hotspot for disease variants causing non-syndromic retinal disease and has a high incidence of variants predicted to significantly alter splicing. We therefore aimed to model exemplar nonsense/frameshift mutations in human induced pluripotent stem cells (iPSCs) to determine their splicing effects and consequently assess the amenability of exon 36 to nonsense-associated altered splicing. We generated two exon 36 mutant iPSC lines: CEP290 ES36 (homozygous: c.4729delinsTT) and CEP290 FS36 (compound heterozygous: c.4730delATinsCAA, c.4719del13) (Figure 4A). All mutations were predicted to be disease causing by Mutation Taster. The ES36 mutation was predicted to alter splicing by Human Splicing Finder (HSF) due to significant alteration of the ESS/ESE ratio, a new acceptor site and activation of a cryptic acceptor site. Of the FS36 mutations, both were predicted to cause nonsense-mediated decay by Mutation Taster and the large 13bp deletion was predicted to impact splicing by HSF (data not shown). RNA analysis of both iPSC lines revealed that ES36 was undergoing skipping of exon 36, whereas FS36 was producing truncated *CEP290* transcript (Figure 4B). Exon 36 skipping in ES36 enabled protein expression to be maintained at a decreased level compared to the wild-type control isogenic iPSCs. In contrast, FS36 had almost complete loss of full-length protein (Figure 4C).

**Figure 4.**
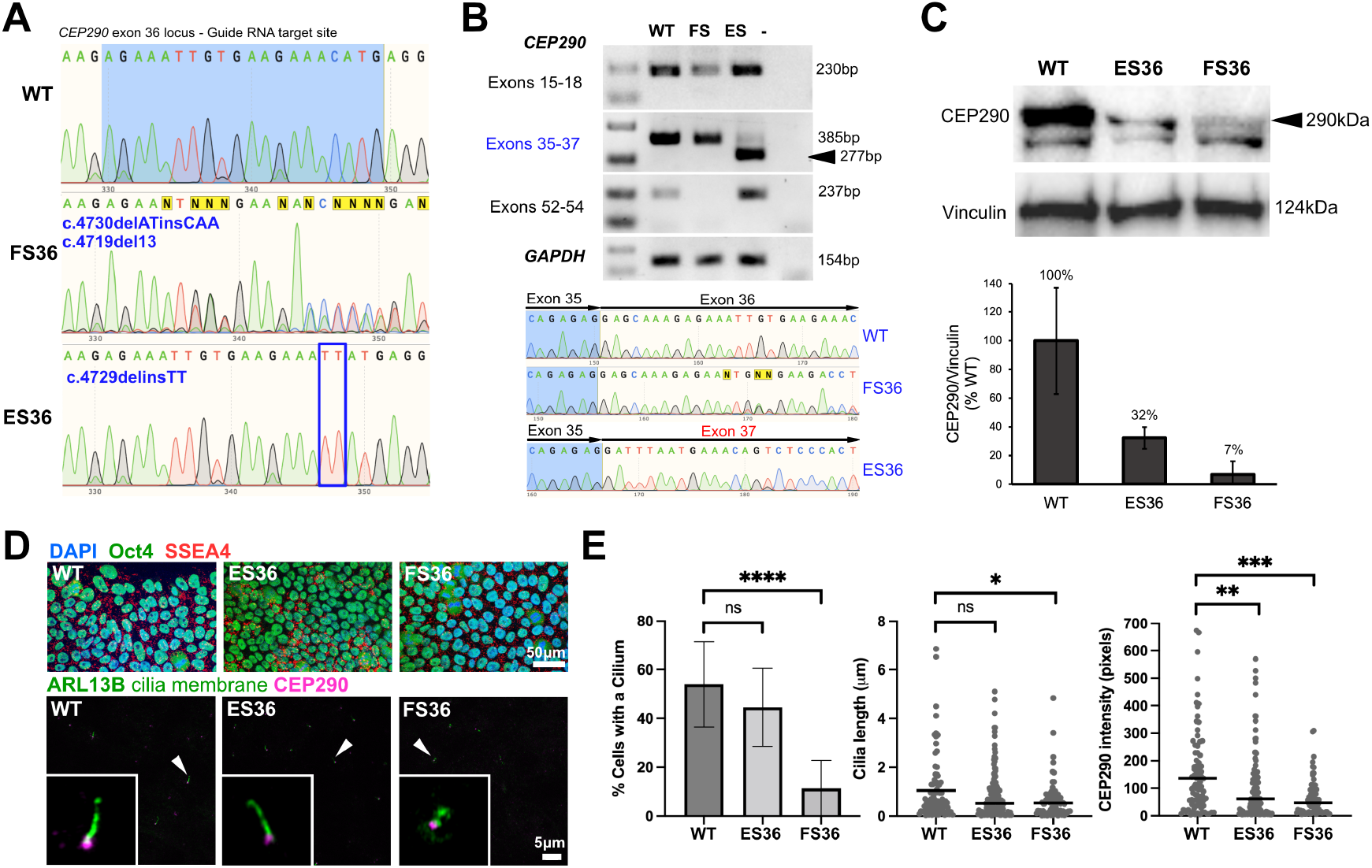
Sequence-specific exon skipping in *CEP290* mutant iPSCs. A. Genotypes presented for CRISPR-edited iPSC lines. ES36 – exon skipped 36, FS36 – frameshift 36, WT – wild type. B – RT-PCR results confirmed exon skipping in ES36 line and truncation in FS36 line in iPSCs. C. CEP290 protein in iPSCs as confirmed by western blot with Vinculin loading control and densitometry analysis results. D. Immunofluorescence staining with pluripotency markers (Oct4 and SSEA4) and cilia markers (ARL13B and CEP290). E. CiliaQ analysis results of ARL13B and CEP290 images from n=3 fields of view, N=3 biological replicates.

Since iPSCs are ciliated, we next assessed the ability of mutant cell lines to undergo ciliogenesis and maintain correct primary ciliary localisation of CEP290 (Figure 4D&E). CEP290 FS36 exhibited a 42% decrease in cilia incidence compared to wild-type control cells (p<0.0001), whereas CEP290 ES36 cells did not have a significant loss of primary cilia. The infrequent primary cilia produced by FS36 cells were significantly shorter (p<0.05) than both the wild-type control and ES36 cells (0.68µm vs 1.05µm and 0.87µm, respectively). In addition, in cells with a primary cilium, the levels of CEP290 localised to the primary cilium were 31.8% for FS36 (p<0.001) and 66.9% for ES36 (p<0.01) compared to wild-type control cilia. We conclude that exon 36 skipping in *CEP290*, can maintain expression of near full-length protein at ∼30% of wild-type levels in iPSCs, and that this is sufficient to maintain normal levels of ciliogenesis and localisation of CEP290, despite the decreased levels of CEP290 expression.

## Discussion

The pleiotropy of *CEP290-*related disorders has only recently gained greater understanding. Recent studies have suggested a role for exon skipping of mutation-containing skiptic exons in ameliorating variant pathogenicity, but this process is likely to be tissue specific. The data presented here suggests a high propensity for *CEP290* variants to cause nonsense-associated altered splicing (NAS), and that NAS is a natural mechanism that attenuates disease phenotype. This is due to enrichment of in-frame exons and the existence of loss-of-function mutations within them, that can cause less severe disease when the exons are spliced out of *CEP290* transcripts. We have also observed that skiptic exon 36 of *CEP290* is a hotspot for LCA mutations and has a high incidence of patient nonsense variants that are predicted to significantly alter splicing. Finally, in exon 36 mutant iPSCs we observed that nonsense-associated altered splicing occurs in a sequence-specific manner and that this does not affect primary cilia formation, in contrast to a frameshift mutant that markedly reduces cilia size and incidence.

There are several case studies of *CEP290* patient blood lymphocytes exhibiting NAS of disease-causing nonsense variants. Additionally, some reports suggest that naturally occurring basal exon skipping (BES) is also a cause of milder disease phenotypes resulting from *CEP290* nonsense variants. Non-canonical BES of *CEP290* exons 6, 8, 10, 18, 32, 41, and 46 have been reported in wild-type skin fibroblasts (9, 10). These studies suggest that some occurrences of milder retinal disease are a result of BES of the affected exon(s) enabling some expression of transcript without a premature stop codon. However, splicing is extremely diverse between cell types, especially in the retina, and it is difficult to draw general conclusions about mechanisms and tissue specificity of BES/NAS from observations made in skin fibroblasts (22, 23).

In structural terms, our *in-silico* analysis predicted 29 coiled coil domains within CEP290. One possibility is that the coiled coils in CEP290 enable it to function as a structural anchor in the primary cilium or to mediate the width of the transition zone by acting as a molecular ruler (3, 24-26). CEP290, by analogy with other large, coiled coil containing proteins, could utilise membrane binding to coordinate the molecular events that occur during target recognition and membrane fusion of ciliary vesicles. This type of behaviour is seen for golgin GCC2/GCC185 (containing 21 coiled coil domains), which is reported to extend for long distances and to act as a tether during Golgi vesicle trafficking and targeting (27). Other large coiled-coil proteins that act as transport vesicle tethers include p115 (USO1 vesicle transport factor) and the endosome tether EEA1 (28). EEA1 undergoes an “entropic collapse” when it binds activated RAB5-GTP, which brings vesicles closer together during membrane fusion (29). *In vitro* biochemical interaction studies of CEP290 protein domains have suggested that cell membrane and microtubule-binding regions at the N- and C-termini regions of CEP290 could mediate a structural role in maintaining the form of the transition zone (4). Consequently, it is plausible that if, for example, a whole coiled coil domain was spliced from *CEP290*, the protein would still be able to form a functional tertiary structure, with fewer coiled coils. For exon 36, which is predicted to be within a link region between two coiled coils in the encoded protein, exon skipping is potentially less deleterious than other exons if it does not affect a coiled coil structure.

The photoreceptor cells in the retina contain specialised primary cilia, with the transition zone (known as the “connecting cilium”) separating the outer and inner segments of the photoreceptor. Here, CEP290 plays a crucial role in regulating the trafficking of proteins essential for light detection (30, 31). As this is a specialised form of primary cilium, it is likely that it is much less able to adapt to any changes in CEP290 availability and/or structure, which may explain retinal phenotypes are a consistent feature of *CEP290*-related disorders.

There are other presentations of *CEP290* disease that may be explained by NAS. Why do some homozygous stop variants cause embryonic lethal disease, whilst others cause less severe yet still syndromic disease? This may be a result of modifier alleles in some cases. But if all nonsense variants resulted in loss-of-function, the effect should be the same. Whereas, if some result in NAS, it may be that for some variants the skipped region is too important to be compatible with functional protein, explaining the differences in phenotype severity.

Most studies functionally characterising *CEP290* variants have been performed in patient fibroblast or lymphocyte cells, cell lines, or mouse models. However, it is apparent that tissue-specific factors play a key role in the pathogenicity of *CEP290* disease. It is therefore crucial going forward that we take advantage of 3D human cellular models of disease organs to study these complex diseases.

Identifying the mechanisms by which *CEP290-*associated disease pathogenesis occurs in retinal cell types, compared with other disease-affected organs, will enable the development of personalised treatments for patients. If canonical exon skipping can be tolerated in disease-affected tissues, this presents an exciting opportunity for therapeutic splice modulation using antisense oligonucleotides for *CEP290*-related disorders.

## Methods

### Exome analysis

The EnsMart function in the Ensembl human genome browser (GRCh38.p13) was used to access and collate datasets for Ensembl gene ID, gene name, physical coordinates for genes and exons, exon phase for predicted splice donor and acceptor sites, and both InterPro and extended Genetic Ontology (GO) terms associated with each gene. The total number of genes (with a GO term annotation) was 19136. The total number of exons (with a unique Ensembl ID) was 576206. The total number of in-phase exons in all three reading frames was 243589 (42.27% of total). Source data files are provided as tab delimited .txt files.

### RNASeq analysis

To analyse native *CEP290* exon usage in disease-relevant organs, short read RNAseq data files were collected for adult human kidney, brain, thyroid, cerebellum, retina and skin, N=3 or 4 (full list, S1). RNASeq FASTQ files were downloaded from the Short Read Archive (https://www.ncbi.nlm.nih.gov/sra) using the SRA Toolkit. All data manipulation and analysis was computed on the University of Leeds ARC4 high performance computer. FASTQ files were trimmed using Trim Galore and quality control checked using FASTQC (https://www.bioinformatics.babraham.ac.uk/projects/fastqc/). Trimmed sequences were aligned using STAR (32), alignment quality was measured by Qualimap(33). Read count files were generated from the aligned BAM files using HTSeq(34) and the latest Ensembl genome file “Homo_sapiens.GRCh38.100.gtf” to define counting bins. Read count tables were analysed for *CEP290* (ENSG00000198707) exon usage using DEXSeq (20), p values were adjusted by Benjamini-Hochberg correction with a false discovery rate of 0.1, strandedness was entered as a confounding factor to account for different library preparation methods. The relative exon usage graph was produced by the DEXSeq programme. This work was undertaken on ARC4, part of the High Performance Computing facilities at the University of Leeds, UK.

### Analysis of CEP290 structure and distribution of nonsense/frameshift variants

The CEP290 coiled-coil structure was analysed using the WAGGAWAGGA server (https://waggawagga.motorprotein.de) (35), using predictions from MARCOIL, MULTICOIL2, NCOILS and PAIRCOIL2. The final coiled-coil prediction shown in Fig 3B was generated by comparison of these predictions and manual adjustment of the predicted coiled-coil register where appropriate. To analyse the distribution of nonsense/frameshift mutations over CEP290 exons in patients, we used patient data from Drivas et al. (2015) and more recent studies (9-11, 17, 36-44). For the analysis we selected patients with exonic nonsense or frameshift mutations in both alleles (145 patients). The data were plotted as a heatmap in MATLAB (see Fig. 3B), showing the frequency of mutations in each exon for each disease.

### Splicing consensus site alignment

To analyse the splice consensus sites of exons with significantly higher usage in the retina (Exons 8, 10, 11, 12, 13, 15, 22 and 39) and those with significantly higher usage in the kidney (Exons 66, 67, 68, 73, 74, 75 and 76), the exon boundary regions were extracted using the table browser tool at UCSC genome browser. For each group, the region 8bp downstream and 8bp upstream of the exon-intron and intron-exon boundaries were aligned using MAFFT^24^ and a sequence logo generated using WebLogo (45, 46).

### Nonsense variant splicing predictions

The NCBI ClinVar database (https://www.ncbi.nlm.nih.gov/clinvar/) was used to identify potential NAS candidates from the list of nonsense variants in *CEP290* (47). The literature was mined to collect available information about patient genotypes and their associated disease. To test the nonsense mutations for potential resulting splicing defects, three independent splice prediction software were used comprising web-based applications (HOT-SKIP and Human Splicing Finder, HSF) and a deep learning-based tool (SpliceAI) (48-50). SpliceAI analysis was done on ARC4 (University of Leeds). The input VCF file was downloaded from ClinVar and reduced to contain all *CEP290* variants using GATK Select Variants (51). The reference genome used was downloaded from NCBI (https://ftp.ncbi.nlm.nih.gov/genomes/refseq/vertebrate_mammalian/Homo_sapiens/latest_assembly_versions/GCF_000001405.39_GRCh38.p13/GCF_000001405.39_GRCh38.p13_genomic.fna.gz) (52). Variants were defined as significantly altering splicing if significant test results were retrieved from ≥ 2 independent splice prediction software.

### iPSC culture

AD2 iPSC lines were a generous gift from Professor Majlinda Lako (Newcastle University, UK) (53). iPSC cells were cultured on 0.083mg/well Matrigel® (Corning) coated 6-well tissue culture plates. Cells were cultured in 2ml mTESR™ Plus medium (StemCell Technologies) at 37°C, 5% CO2. Medium changes were made daily Sun-Thurs, with 4ml medium given on Fridays. Cells were defrosted in a 37°C water bath and transferred to a fresh 50ml falcon tube. 6ml room temperature medium containing 10µM Y-27632 (StemCell Technologies) was added dropwise to cells. Cells were centrifuged (200*g*, 5min) to remove dimethyl sulphoxide (DMSO). Cells were resuspended in 2ml 10µM Y-27632 media and seeded into two wells of a 6-well Matrigel® plate, already containing 1ml 10µM Y-27632 media at 37°C. Cells were passaged after 48hrs. To passage confluent cells, medium was removed and cells were washed in 2ml Ca^2+^/Mg^2+^ free phosphate-buffered saline (PBS) and treated for 45sec with 1ml ReLeSR™ (StemCell Technologies). ReLeSR was immediately removed and cells were incubated at 37°C for 7 minutes. 1ml fresh medium was applied to cells and the plate gently tapped to resuspend cells. The cell suspension was added to fresh medium (volume dependent on passage ratio) and mixed by pipetting up and down twice, slowly. Cells were passaged every three days, with a maximum ratio of 1:8.

### CRISPR-Cas9 gene editing of CEP290 exon 36 in iPSCs

To model specific *CEP290* variants in iPSCs, four guide RNAs were first chosen from Benchling targeting exon 36 (54). These were selected based on high efficiency and off-target scores. Single stranded DNA oligonucleotides of each guide RNA were cloned into PX458 (Cas9-GFP, Addgene #48138) and cloning was confirmed by Sanger sequencing (Genewiz, UK).

To select one guide RNA for each target site to take forward for editing in iPSCs, the most efficient guides were identified. 10^4^ HEK293 cells were seeded per well into a 96-well plate in 10% v/v FBS in DMEM and left to grow for 72 hours in a 37°C, 5% CO_2_ incubator. 200ng miniprepped gRNA-PX458 plasmid was mixed with 25μl Opti-MEM™ and incubated for 5 minutes. 0.6μl Lipofectamine 2000 was added to 25μl Opti-MEM™ and incubated for 5 minutes. The Lipofectamine suspension was added to the DNA solution and pipette mixed. The transfection complexes were incubated for 20 minutes at room temperature. After this, medium on the HEK293 cells was replaced for 50µl 10% FBS v/v DMEM. 50µl transfection complexes were added to each well dropwise and incubated for 5 hours, then medium changed for 200µl fresh 10% v/v FBS in DMEM. Transfected cells were incubated for 72 hours at 37°C, 5% CO_2_.

To determine the efficiency of each guide, DNA analysis was performed. DNA was extracted from each transfected well using DirectPCR Lysis Reagent (Cell) (Viagen Biotech) according to the manufacturer’s instructions. HotShot Diamond (Clent Life Science) PCR reactions were done to amplify around each target site within *CEP290* and the products were cleaned up enzymatically with ExoSAP-IT™ (Thermo Fisher Scientific), then sent for Sanger sequencing (Genewiz). Sequences were analysed using TIDE (https://tide.nki.nl/) and the guide RNA with the highest % edited sequences taken forward (55).

iPSCs were grown to near confluency in a 6-well plate, washed briefly in Mg/Ca^2+^-free PBS, then incubated in 0.75ml of 0.75X TrypLE™ Express Enzyme (Thermo Fisher Scientific) at 37°C, 5% CO_2_ for 10 minutes to achieve a single cell suspension. 1.5ml mTESR with 10µM Y-27632 was added to each cell suspension to halt TrypLE™ activity. Cells were diluted to 7×10^5^cells in 1ml media and then centrifuged for 5mins, 200g. Supernatant was removed, and cells were resuspended in 100µl Nucleofector® Solution 2 (Lonza). 2µg maxiprepped gRNA-PX458 plasmid was added to the reaction and pipette mixed gently. The reaction mix was transferred to a Lonza certified cuvette, then electroporated using an Amaxa® Nucleofector® II (Lonza), setting A-023. 500µl mTESR with 10µM Y-27632 was added to the electroporated cells and then a thin Pasteur pipette used to carefully transfer the cells from the cuvette and add them dropwise to a warm Matrigel® (Corning)-prepared 24-well plate containing 500µl mTESR with 10µM Y-27632. Electroporated cells were then incubated for 48 hours at 37°C, 5% CO_2_.

To expand clonal populations from positively transfected cells, flow assisted cell sorting (FACS) was used. Two 96-well plates of GFP-positive single cells were prepared and monitored over 14 days. Colonies were identified using an Operetta High Content Imaging System (PerkinElmer) and passaged 1:2 into two wells of a fresh Matrigel®-prepared 96-well plate. One well was used for continued expansion, the other was taken for DNA analysis as before. Sequence electropherograms were analysed in Snapgene Viewer and using EMBOSS-Needle alignment tool to identify genomic edits.

### RT-PCR analysis

To look at variable transcript expression arising from *CEP290* variants, a set of RT-PCR experiments were performed. RNA was extracted from confluent iPSCs and organoids using Trizol® as per Manufacturer’s instructions. Turbo DNAse treated RNA samples were cleaned up using Zymo RNA Clean and Concentrator™-5 kit according to the manufacturer’s instructions. Then cDNA was synthesised using SuperScript™ II Reverse Transcriptase (Thermo Fisher) according to the manufacturer’s instructions with 500ng RNA. cDNA was diluted 1:50 in nuclease-free water for RT-PCR reactions. PCR reactions were set up with 3µl HotShot Diamond, 1µl 10µM F&R primer pair, 4µl nuclease-free water, and 2µl diluted cDNA or 2µl nuclease-free water (negative control). RT-PCR products were analysed by gel electrophoresis on a 2% agarose gel, stained with Midori Green (120V, 35mins) and imaged with a ChemiDoc™ MP Imaging System (Bio-Rad Laboratories). Band density analysis was carried out using Image Lab Software (Bio-Rad Laboratories), normalised against *GAPDH* RT-PCR bands and student’s t test carried out.

### Western blot analysis

Western blotting was used to compare CEP290 protein expression levels between wild-type control and CEP290 FS36 and ES36 lines. Cells were washed in 2ml ice cold Ca^2+^/Mg^+^ PBS. 200µl ice cold Pierce IP Lysis Buffer (Invitrogen), with Halt Protease and Phosphatase Inhibitor Cocktails added at 1:100 dilution (Thermo Fisher Scientific), was added to each well containing confluent cells. The plate was incubated on ice for 5 minutes, pipette mixing intermittently. Lysed cells were transferred to cold 1.5ml tubes and snap frozen in liquid LN_2_ before storage at -80°C. Lysed cells were defrosted on a rotator at 4°C for 30 minutes, then centrifuged at 13000 x g for 10 minutes at 4°C to remove insoluble material. The supernatant containing soluble protein material was transferred to a fresh cold 1.5ml tube and protein concentration determined by BSA assay. 30µg cell lysate was diluted in Pierce IP lysis buffer up to 18.75µl total volume on ice, followed by the addition of 6.25µl 20% (v/v) β-mercaptoethanol 4x LDS (final concentrations 5%, 1X, respectively) to a final volume 25µl. Proteins were denatured for 10 minutes at 95°C and then loaded on a NuPAGE™ 3-8% Tris-Acetate 1.0mm 12-well Protein Gel (Invitrogen™, Thermo Fisher Scientific) alongside HiMark™ Pre-stained Protein Standard (Invitrogen™, Thermo Fisher Scientific) and run for 2hr15mins at 150V, then transferred to a PVDF membrane overnight at 10V.

Membranes were washed for 5 minutes in PBS-T (containing 0.1% Tween-20), then transferred to a 50ml tube and blocked for 1 hour in 3% BSA w/v PBS-T on a roller at room temperature (RT). The BSA solution was removed from the tube and replaced with primary antibody diluted in 3% BSA w/v PBS-T (rabbit anti-CEP290, Novus Biologicals, 1:2000; mouse anti-Vinculin, 1:5000, Sigma Aldrich) and incubated on roller overnight at 4°C or for 1 hour 30 minutes at RT. Membranes were washed in PBS-T for 5x 2 minutes and 1x 20 minutes, followed by incubation in secondary antibody diluted in 3% BSA w/v PBS-T on roller for 1hr or 15 minutes at RT (porcine anti-rabbit HRP, 1:5000; Streptavidin-HRP, 1:1000; goat anti-mouse HRP, 1:5000), before removing and washing as before. Bands were detected using ECL substrate (SuperSignal™ West Femto Maximum Sensitivity Substrate, Thermo Fisher Scientific), following the manufacturer’s instructions, and imaged with a ChemiDoc™ MP Imaging System (Bio-Rad Laboratories). Densitometric analysis was carried out using Image Lab Software (Bio-Rad Laboratories), normalisation against Vinculin bands and Student’s t test done in Excel, and graphs generated in Prism (GraphPad).

### Immunocytochemistry

Cells were seeded on Matrigel^®^-coated coverslips (Academy Science, NPC16/13). When confluent, medium was removed and cells washed in Ca^2+^/Mg^2+^-containing PBS. Cells were either fixed in 1ml 4% paraformaldehyde (PFA) (room temperature, 20min) or 1ml 100% ice-cold MeOH (−20°C, 5mins). PFA-fixed cells were permeabilised in 1ml 0.1% (v/v) Triton^™^-X-100 (Sigma Aldrich) in PBS on a rocker (5mins). Cells were blocked in 3% normal goat serum (NGS) or normal donkey serum (NDS), 0.05% Triton^™^-X-100 [v/v] PBS on a rocker. Antibodies were diluted in blocking solution and centrifuged (16000 x g, 5mins), to remove particulates. Coverslips were inverted onto 30µl primary antibody dilutions in a humid closed chamber and incubated at 4°C overnight (mouse anti-SSEA4 conjugated with Alexa Fluor^®^555, BD Biosciences, 1:10; goat anti-OCT4, R&D Biosystems, 1:10; mouse anti-CEP290, generous gift from Dr Ciaran Morrison, National University of Ireland, Galway; 1:100; rabbit anti-ARL13B, Proteintech^®^, 1:2000). Coverslips were then washed five times for 2min in 0.05% Triton-X-100 [w/v] PBS on a rocker. Coverslips were inverted onto 30µl secondary antibody dilutions in a humid chamber and incubated for 1.5hrs at room temperature in the dark (donkey anti-goat-IgG AlexaFluor488, 1:2000; Donkey anti-rabbit-IgG AlexaFlour488, 1:2000; Donkey anti-mouse-IgG AlexaFluor568, 1:2000, all Thermo Fisher Scientific). Previous wash steps were repeated, final incubation in 30µl DAPI stain (Thermo Fisher Scientific) for 20mins. Two final washes were carried out, with a final wash in dH_2_O. Coverslips were mounted on microscope slides with ProLong^™^ Gold Antifade Mountant (Thermo Fisher Scientific). Cells were imaged using a Nikon A1R confocal microscope or Zeiss LSM880 inverted confocal microscope with Airyscan.

Images were processed in Fiji Image J. Cilia length, incidence, and CEP290 localisation were analysed using the ImageJ plugin CiliaQ (56). Statistical analysis was done in Excel (Z score – standard error of the mean or Student’s t test), and graphs generated in Prism (GraphPad).

## Data Availability

All data produced in the present study are available upon reasonable request to the authors

## Acknowledgements

We thank colleagues at the LIMR Flow and Imaging Facility, University of Leeds, for help with imaging and microscopy. This work was undertaken on ARC4, part of the High-Performance Computing facilities at the University of Leeds, UK. We are grateful to Prof. Ciaran Morrison (National University of Ireland Galway) for the gift of the CEP290 mouse monoclonal antibody. R.D.T is supported by post-graduate studentship funded by University of Leeds alumnus donation from Mr. Clive Summerhayes. C.A.J acknowledges support from Medical Research Council project grants MR/M000532/1 and MR/T017503/1. J.A.P. is supported by a UKRI Future Leaders Fellowship (MR/T02044X/1).

