## Supplementary Tables for "The role of alternative splicing in CEP290-related disease pathogenesis"

### Supplementary Table 1

| Chr12 GRCH38 | Exon | Variant | Protein | Class | ClinVar listed phenotypes | Published Phenotype | Allele 1 | Allele 2 | Source |
|---|---|---|---|---|---|---|---|---|---|
| 88050409 | 53 | c.7153del | p.Lys2384_Ile2385insTer | P | JBTS, MKS, NPHP | N/A | | | |
| 88053703 | 52 | c.7073_7077dup | p.His2360Ter | P | NPHP, MKS, JBTS | N/A | | | |
| 88053733 | 52 | c.7048C>T | p.Gln2350Ter | LP | not provided | LCA | c.7048C>T, p.Gln2350Ter | c.4029+1G>A | https://www.ajo.com/article/S0002-9394(19)30551-3/fulltext |
| | | | | | | Retinal Dystrophy | c.7048C>T, p.Gln2350Ter | c.1711G>A, p.Gly571Arg | https://www.ncbi.nlm.nih.gov/pmc/articles/PMC5565704/ |
| 88055597 | 50 | c.6939C>A | p.Tyr2313Ter | P | JBTS | JBTS | c.6939C>A, p.Tyr2313Ter | c.1219_1220delAT, p.Met407GlufsTer14 | https://www.ncbi.nlm.nih.gov/pmc/articles/PMC5082428/ |
| 88055644 | 50 | c.6892C>T | p.Gln2298Ter | LP | Retinal Dystrophy | N/A | | | |
| 88055665 | 50 | c.6871C>T | p.Gln2291Ter | LP | LCA | LCA | c.6871C>T, p.Gln2291Ter | c.180+1G>A | https://www.ncbi.nlm.nih.gov/pmc/articles/PMC6362094/ |
| 88055700 | 50 | c.6836T>A | p.Leu2279Ter | P/LP | Retinal Dystrophy, JBTS, MKS, NPHP | N/A | | | |
| 88058868 | 49 | c.6798G>A | p.Trp2266Ter | P | JBTS, NPHP, MKS | Episodic Ataxia | c.6798G>A, p. Trp2266Ter | c.2174A>C, p.Glu725Ala | https://onlinelibrary.wiley.com/doi/full/10.1002/mdc3.12872 |
| 88058869 | 49 | c.6797G>A | p.Trp2266Ter | P | not provided | N/A | | | |
| 88059909 | 48 | c.6634G>T | p.Glu2212Ter | P | NPHP, MKS, JBTS | N/A | | | |
| 88060988 | 47 | c.6364A>T | p.Arg2122Ter | P | not provided | N/A | | | |
| 88062718 | 46 | c.6331C>T | p.Gln2111Ter | unknown | not provided | JSRD | c.6331C>T, p.Gln2111Ter | ? | https://www.nature.com/articles/ng1786/tables/1 |
| 88068585 | 44 | c.6072C>A | p.Tyr2024Ter | P | JBTS | JSRD | c.6072C>A, p.Tyr2024Ter | c.7321dupCTCT, p.Ley2440fsTer2456 | https://www.cell.com/ajhg/fulltext/S0002-9297(07)62820-1 |
| | | | | | | JSRD | c.6072C>A, p.Tyr2024Ter | ? | https://www.ncbi.nlm.nih.gov/pmc/articles/PMC5682233/ |
| 88068626 | 44 | c.6031C>T | p.Arg2011Ter | P | not provided | JSRD | c.6031C>T, p.Arg2011Ter | c.1657_1666delA, p.Leu552fsTer572 | https://www.cell.com/ajhg/fulltext/S0002-9297(07)62820-1 |

| | | | | | | | | | |
|---|---|---|---|---|---|---|---|---|---|
| | | | | | | RP | c.6031C>T, p.Arg2011Ter | ? | https://www.aaojournal.org/article/S0161-6420(19)30438-5/fulltext#supplementaryMaterial |
| 88071373 | 43 | c.5932C>T | p.Arg1978Ter | P | JBTS | SLS | c.5932C>T, p.Arg1978Ter | ? | https://www.ncbi.nlm.nih.gov/pmc/articles/PMC2597962/ |
| | | | | | | JSRD | c.5932C>T, p.Arg1978Ter | c.164_167delCTCA, p.Thr55SerfsTer3 | https://www.ncbi.nlm.nih.gov/pmc/articles/PMC5082428/ |
| 88071833 | 42 | c.5803G>T | p.Glu1935Ter | P | LCA | N/A | | | |
| 88071848 | 42 | c.5788A>T | p.Lys1930Ter | P | LCA | JSRD | c.5788A>T, p.Lys1930Ter | c.6012-12A>T | https://www.nature.com/articles/jhg2012117 |
| 88071860 | 42 | c.5776C>T | p.Arg1926Ter | P/LP | NPHP, MKS, JBTS | LCA | c.5776C>T, p.Arg1926Ter | ? | https://www.ajo.com/article/S0002-9394(07)00767-2/fulltext |
| 88071890 | 42 | c.5745dup | p.Lys1916Ter | P | NPHP, MKS, JBTS | N/A | | | |
| 88077224 | 41 | c.5707A>T | p.Lys1903Ter | P | NPHP | BBS | **c.5707A>T, p.Lys1903Ter** | **c.5707A>T, p.Lys1903Ter** | |
| 88077227 | 41 | c.5704G>T | p.Glu1902Ter | P | BBS, JBTS | JSRD | **c.5704G>T, p.Glu1902Ter** | **c.5704G>T, p.Glu1902Ter** | https://pubmed.ncbi.nlm.nih.gov/26092869/ https://pubmed.ncbi.nlm.nih.gov/31464256/ |
| 88077263 | 41 | c.5668G>T | p.Gly1890Ter | P | SLS, JBTS, Retinal Dystrophy, MKS, LCA, RP, MKS | LCA | c.5668G>T, p.Gly1890Ter | c.2991+1665A>G, p.Cys998Ter | https://www.ajo.com/article/S0002-9394(19)30551-3/fulltext |
| | | | | | | JSRD | **c.5668G>T, p.Gly1890Ter** | **c.5668G>T, p.Gly1890Ter** | https://www.nature.com/articles/ng1786/tables/1 |
| | | | | | | JSRD | **c.5668G>T, p.Gly1890Ter** | **c.5668G>T, p.Gly1890Ter** | https://jmg.bmj.com/content/53/11/761.long |
| | | | | | | JSRD | c.5668G>T, p.Gly1890Ter | c.1189G>A, p.Gly397Ser | https://www.ncbi.nlm.nih.gov/pmc/articles/PMC4037742/ |
| | | | | | | JSRD | c.5668G>T, p.Gly1890Ter | c.4656del1, p.Lys1552fsTer1923 | https://www.ncbi.nlm.nih.gov/pmc/articles/PMC4037742/ |
| | | | | | | RP | **c.5668G>T, p.Gly1890Ter** | **c.5668G>T, p.Gly1890Ter** | https://www.ncbi.nlm.nih.gov/pmc/articles/PMC6362094/ |
| 88079112 | 39 | c.5344C>T | p.Arg1782Ter | P | JBTS, NPHP, MKS, LCA, BBS, SLS | LCA | c.5344C>T, p.Arg1782Ter | c.2991+1665A>G, p.Cys998Ter | https://www.ncbi.nlm.nih.gov/pmc/articles/PMC3048164/ |
| | | | | | | JSRD | c.5344C>T, p.Arg1782Ter | c.1666dupA, p.Ile556AsnfsTer20 | https://pubmed.ncbi.nlm.nih.gov/26092869/ |
| 88080196 | 38 | c.5212G>T | p.Glu1738Ter | P | NPHP, MKS, JBTS | N/A | | | |

| | | | | | | | | | |
|---|---|---|---|---|---|---|---|---|---|
| 88080226 | 38 | c.5182G>T | p.Glu1728Ter | P | NPHP, MKS, JBTS | SECORD | c.5182G>T, p.Glu1782Ter | c.508A>T, p.Lys170Ter | https://www.ncbi.nlm.nih.gov/pmc/articles/PMC5565704/ |
| 88083083 | 37 | c.4960C>T | p.Gln1654Ter | P | LCA | LCA | c.4960C>T, p.Gln1654Ter | c.2991+1665A>G, p.Cys998Ter | https://www.nature.com/articles/ejhg20179#MOESM236 |
| 88083161 | 37 | c.4882C>T | p.Gln1628Ter | P | JBTS, SLS, BBS, MKS, LCA, NPHP | LCA | c.4882C>T, p.Gln1628Ter | c.2991+1665A>G, p.Cys998Ter | https://www.ajo.com/article/S0002-9394(19)30551-3/fulltext |
| | | | | | | JSRD | c.4882C>T, p.Gln1628Ter | c.5941G>T, p.Glu1981Ter | https://www.ncbi.nlm.nih.gov/pmc/articles/PMC4037742/#SD1 |
| | | | | | | JSRD | c.4882C>T, p.Gln1628Ter | c.5611_5614delCAAA, p.Gln1871ValfsTer2 | https://pubmed.ncbi.nlm.nih.gov/26092869/ |
| 88083848 | 36 | c.4811G>A | p.Trp1604Ter | P | JBTS | JSRD | **c.4811G>A, p.Trp1604Ter** | **c.4811G>A, p.Trp1604Ter** | https://www.ncbi.nlm.nih.gov/pmc/articles/PMC5126998/ |
| 88083858 | 36 | c.4801C>T | p.Gln1601Ter | LP | Retinal Dystrophy | LCA | c.4801C>T, p.Gln1601Ter | c.2991+1665A>G, p.Cys998Ter | https://jmg.bmj.com/content/53/11/761.long |
| 88083888 | 36 | c.4771C>T | p.Gln1591Ter | P | LCA | JSRD | c.4771C>T, p.Gln1591Ter | ? | https://www.nature.com/articles/ng1786/tables/1 |
| | | | | | | LCA | **c.4771C>T, p.Gln1591Ter** | **c.4771C>T, p.Gln1591Ter** | https://iovs.arvojournals.org/article.aspx?articleid=2189538 |
| 88083927 | 36 | c.4732G>T | p.Glu1578Ter | not provided | not provided | JSRD | **c.4732G>T, p.Glu1578Ter** | **c.4732G>T, p.Glu1578Ter** | https://www.nature.com/articles/ng1805#MOESM1 |
| | | | | | | LCA | c.4732G>T, p.Glu1578Ter | c.3012delA, p.Lys1004fs | https://www.ncbi.nlm.nih.gov/pmc/articles/PMC5664130/ |
| 88083936 | 36 | c.4723A>T | p.Lys1575Ter | P | JBTS, NPHP, LCA, MKS, SLS | LCA | c.4723A>T, p.Lys1575Ter | c.2991+1665A>G, p.Cys998Ter | https://www.ajo.com/article/S0002-9394(19)30551-3/fulltext |
| | | | | | | LCA | **c.4723A>T, p.Lys1575Ter** | **c.4723A>T, p.Lys1575Ter** | https://www.ncbi.nlm.nih.gov/pmc/articles/PMC3048164/ |
| | | | | | | LCA | c.4723A>T, p.Lys1575Ter | c.4696G>C, p.Ala1566Pro | https://www.ncbi.nlm.nih.gov/pmc/articles/PMC3048164/ |
| | | | | | | LCA | c.4723A>T, p.Lys1575Ter | c.1709C>G, p.Ser570Ter | https://onlinelibrary.wiley.com/doi/epdf/10.1002/humu.9485 |
| | | | | | | SLS | c.4723A>T, p.Lys1575Ter | c.1984C>T, p.Gln662Ter | https://www.ncbi.nlm.nih.gov/pmc/articles/PMC4643834/ |
| | | | | | | Liver fibrosis, intellectual disability, nystagmus, strabismus | c.4723A>T, p.Lys1575Ter | c.1936C>T, p.Gln646Ter | https://www.ncbi.nlm.nih.gov/pmc/articles/PMC4643834/ |
| | | | | | | SLS | c.4723A>T, p.Lys1575Ter | c.4393C>T, p.Arg1465Ter | https://www.ncbi.nlm.nih.gov/pmc/articles/PMC3048164/ |

| | | | | | | | | | |
|---|---|---|---|---|---|---|---|---|---|
| | | | | | | JSRD | c.4723A>T, p.Lys1575Ter | c.4393C>T, p.Arg1465Ter | https://www.ncbi.nlm.nih.gov/pmc/articles/PMC1950920/ |
| | | | | | | JSRD | c.4522C>T, p.Arg1508Ter | c.4723A>T, p.Lys1575Ter | https://www.ncbi.nlm.nih.gov/pmc/articles/PMC5082428/ |
| | | | | | | SLS | c.4723A>T, p.Lys1575Ter | c.1987A>T, p.Gln663Ter | https://www.researchgate.net/publication/233788878_High-throughput_mutation_analysis_in_patients_with_a_nephronophthisis_associated_ciliopathy_applying_multiplexed_barcoded_array-based_PCR |
| | | | | | | SLS | c.4723A>T, p.Lys1575Ter | c.3802C>T, p.Gln1268Ter | https://www.researchgate.net/publication/233788878_High-throughput_mutation_analysis_in_patients_with_a_nephronophthisis_associated_ciliopathy_applying_multiplexed_barcoded_array-based_PCR |
| | | | | | | SLS | c.4723A>T, p.Lys1575Ter | c.1189+1G>A | https://www.ncbi.nlm.nih.gov/pmc/articles/PMC4643834/ |
| | | | | | | Retinal Dystrophy | c.4723A>T, p.Lys1575Ter | c.6599delA, p.Glu2200del1gA | https://www.ncbi.nlm.nih.gov/pmc/articles/PMC5565704/ |
| 88084768 | 35 | c.4522C>T | p.Arg1508Ter | P | JBTS, MKS, NPHP | JSRD | c.4522C>T, p.Arg1508Ter | c.4723A>T, p.Lys1575Ter | https://www.ncbi.nlm.nih.gov/pmc/articles/PMC5082428/ |
| 88084800 | 35 | c.4490C>A | p.Ser1497Ter | LP | Retinal Dystrophy | N/A | | | |
| 88086083 | 34 | c.4393C>T | p.Arg1465Ter | P | JBTS, LCA, NPHP, MKS, SLS, BBS | LCA | c.4393C>T, p.Arg1465Ter | c.2991+1665A>G, p.Cys998Ter | https://www.ncbi.nlm.nih.gov/pmc/articles/PMC3048164/ |
| | | | | | | JSRD | c.4393C>T, p.Arg1465Ter | c.4723A>T, p.Lys1575Ter | https://www.ncbi.nlm.nih.gov/pmc/articles/PMC1950920/ |
| 88086415 | 33 | c.4276_4277del | p.Gln1425_Asn1426insTer | P | JBTS | N/A | | | |
| 88086450 | 33 | c.4243G>T | p.Glu1415Ter | LP | MKS, LCA, JBTS, BBS, SLS | JSRD | c.4243G>T, p.Glu1415Ter | ? | https://pubmed.ncbi.nlm.nih.gov/29754767/ |
| 88087788 | 32 | c.4186C>T | p.Gln1396Ter | P | JBTS, NPHP, MKS | N/A | | | |
| 88087934 | 32 | c.4040G>A | p.Trp1347Ter | P | NPHP, JBTS, MKS | RP | c.4040G>A, p.Trp1347Ter | c.3104-2delA | https://www.spandidos-publications.com/mmr/11/3/1827 |
| 88089118 | 31 | c.3943G>T | p.Glu1315Ter | P | not provided | N/A | | | |
| 88089157 | 31 | c.3904C>T | p.Gln1302Ter | P | JBTS, LCA | LCA | c.3904C>T, p.Gln1302Ter | c.6869_6870insA, p.Asn2290LysfsTer6 | https://www.jmdjournal.org/article/S1525-1578(14)00176-7/fulltext |

| | | | | | | | | | |
|---|---|---|---|---|---|---|---|---|---|
| | | | | | | LCA | c.3904C>T, p.Gln1302Ter | c.4661_4663del, p.Glu1544del and c.-1G>A | https://www.ncbi.nlm.nih.gov/pmc/articles/PMC7043639/ |
| | | | | | | JSRD | c.3904C>T, p.Gln1302Ter | c.1666delA, p.Ile556Phefs*17 | https://www.ncbi.nlm.nih.gov/pmc/articles/PMC5082428/ |
| 88089166 | 31 | c.3894dup | p.Lys1299Ter | P/LP | MKS, NPHP, JBTS | JSRD | **c.3894dup, p.Lys1299Ter** | **c.3894dup, p.Lys1299Ter** | https://www.nature.com/articles/ejhg2016146#Sec17 |
| 88089247 | 31 | c.3814C>T | p.Arg1272Ter | P | NPHP, MKS, JBTS | LCA | c.3814C>T, p.Arg1272Ter | c.2991+1665A>G, p.Cys998Ter | https://linkinghub.elsevier.com/retrieve/pii/S0002-9297(07)62755-4 |
| 88089259 | 31 | c.3802C>T | p.Gln1268Ter | P | NPHP, MKS, JBTS | SLS | c.3802C>T, p.Gln1268Ter | c.4723A>T, p.Lys1575Ter | https://www.researchgate.net/publication/233788878_High-throughput_mutation_analysis_in_patients_with_a_nephronophthisis_associated_ciliopathy_applying_multiplexed_barcoded_array-based_PCR |
| 88089468 | 31 | c.3593C>A | p.Ser1198Ter | P | NPHP, MKS, JBTS | N/A | | | |
| 88093839 | 28 | c.3420T>G | p.Tyr1080Ter | P | JBTS, NPHP, MKS | N/A | | | |
| 88093859 | 28 | c.3220G>T | p.Glu1074Ter | LP | Retinal Dystrophy | N/A | | | |
| 88093873 | 28 | c.3205_3206delinsTG | p.Glu1069Ter | P | NPHP, MKS, JBTS | N/A | | | |
| 88093888 | 28 | c.3190del | p.Glu1063_Met1064insTer | P | NPHP, MKS, JBTS | MKS | **c.3190del, p.Met1064Ter** | **c.3190del, p.Met1064Ter** | https://academic.oup.com/hmg/article/24/18/5211/687840 |
| 88093903 | 28 | c.3175del | p.Lys1058_Ile1059insTer | LP | MKS | MKS | **c.3175del, p.Ile1059Ter** | **c.3175del, p.Ile1059Ter** | https://www.ncbi.nlm.nih.gov/pmc/articles/PMC2718326/ |
| 88093955 | 28 | c.3123dup | p.Lys1042Ter | P | not provided | N/A | | | |
| 88096894 | 27 | c.3097A>T | p.Lys1033Ter | LP | not provided | LCA | c.3097A>T, p.Lys1033Ter | c.1910-11T>C | https://pubmed.ncbi.nlm.nih.gov/28714225/ |
| 88102888 | 26 | c.2941C>T | p.Gln981Ter | P | JBTS?? | N/A | | | |
| 88102918 | 26 | c.2911G>T | p.Glu971Ter | P | not provided | N/A | | | |
| 88106770 | 25 | c.2722C>T | p.Arg908Ter | P | JBTS, cone-rod dystrophy | Cone-Rod Dystrophy | **c.2722C>T, p.Arg908Ter** | **c.2722C>T, p.Arg908Ter** | https://www.ncbi.nlm.nih.gov/pmc/articles/PMC6362094/ |
| 88106784 | 25 | c.2708T>G | p.Leu903Ter | VUS | not provided | N/A | | | |
| 88106887 | 25 | c.2605C>T | p.Gln869Ter | P | JBTS, MKS, NPHP | Syndromic retinal dystrophy | **c.2605C>T, p.Gln869Ter** | **c.2605C>T, p.Gln869Ter** | https://www.ncbi.nlm.nih.gov/pmc/articles/PMC6978239/ |
| 88111218 | 22 | c.2351T>A | p.Leu784Ter | LP | not provided | N/A | | | |

| | | | | | | | | | |
|---|---|---|---|---|---|---|---|---|---|
| 88111318 | 22 | c.2251C>T | p.Arg751Ter | P | JBTS | MKS | c.2251C>T, p.Arg751Ter | c.4864_4865delAA, p.Ser820PhefsTer9 | https://www.ncbi.nlm.nih.gov/pmc/articles/PMC4643834/ |
| | | | | | | JSRD | c.2251C>T, p.Arg751Ter | c.6869delA, p.Asn2290fsTer2300 | https://jasn.asnjournals.org/content/18/5/1566.long |
| 88111320 | 22 | c.2249T>G | p.Leu750Ter | P | LCA | LCA | c.2249T>G, p.Leu750Ter | c.2991+1665A>G, p.Cys998Ter | https://www.cell.com/ajhg/fulltext/S0002-9297(07)62755-4 |
| | | | | | | Retinal Dystrophy | c.2249T>G, p.Leu750Ter | c.4063C>T, p.Leu750Ter | https://www.ncbi.nlm.nih.gov/pmc/articles/PMC5565704/ |
| 88114488 | 20 | c.1984C>T | p.Gln662Ter | P | JBTS, NPHP, MKS, LCA, RP | Retinal Dystrophy | c.1984C>T, p.Gln662Ter | c.4723A>T, p.Lys1575Ter | https://www.ncbi.nlm.nih.gov/pmc/articles/PMC4643834/ |
| | | | | | | LCA | c.1984C>T, p.Gln662Ter | c.5587-1G>C | https://www.ncbi.nlm.nih.gov/pmc/articles/PMC6362094/ |
| | | | | | | RP | **c.1984C>T, p.Gln662Ter** | **c.1984C>T, p.Gln662Ter** | https://www.ncbi.nlm.nih.gov/pmc/articles/PMC6362094/ |
| 88114536 | 20 | c.1936C>T | p.Gln646Ter | P | NPHP, JBTS, MKS, Retinal Dystrophy | LCA | c.1936C>T, p.Gln646Ter | c.2991+1665A>G, p.Cys998Ter | https://www.ajo.com/article/S0002-9394(19)30551-3/fulltext |
| | | | | | | LCA | c.1936C>T, p.Gln646Ter | c.6604delA, p.Ile2203LeufsTer23 | https://onlinelibrary.wiley.com/doi/epdf/10.1002/humu.9485 |
| | | | | | | Liver fibrosis, intellectual disability, nystagmus, strabismus | c.1936C>T, p.Gln646Ter | c.4723A>T, p.Lys1575Ter | https://www.ncbi.nlm.nih.gov/pmc/articles/PMC4643834/ |
| 88115149 | 19 | c.1858G>T | p.Glu620Ter | P | NPHP, MKS, JBTS | N/A | | | |
| 88117076 | 18 | c.1781T>A | p.Leu594Ter | P | Retinal Dystrophy | LCA | c.1781T>A, p.Leu594Ter | c.2991+1665A>G, p.Cys998Ter | https://www.ncbi.nlm.nih.gov/pmc/articles/PMC5106339/#SM1 |
| 88118485 | 17 | c.1709C>G | p.Ser570Ter | LP | not provided | LCA | c.1709C>G, p.Ser570Ter | c.4723A>T, p.Lys1575Ter | https://onlinelibrary.wiley.com/doi/epdf/10.1002/humu.9485 |
| 88118549 | 17 | c.1645C>T | p.Arg549Ter | P | JBTS, NPHP, MKS | LCA | c.1645C>T, p.Arg549Ter | c.4661_4663del, p.Glu1544del | https://www.ncbi.nlm.nih.gov/pmc/articles/PMC3283211/ |
| | | | | | | JSRD | c.1645C>T, p.Arg549Ter | c.5649insA, Lys1884fsTer1906 | https://jasn.asnjournals.org/content/18/5/1566.long#T1 |
| | | | | | | JSRD | c.1645C>T, p.Arg549Ter | c.5643_5644insA, p.Lys1882ins1gttA | https://www.ncbi.nlm.nih.gov/pmc/articles/PMC5565704/ |
| 88118567 | 17 | c.1627G>T | p.Glu543Ter | P | JBTS, MKS, NPHP | LCA | c.1627G>T, p.Glu543Ter | c..367C>T, p.Gln123Ter | https://iovs.arvojournals.org/article.aspx?articleid=2624456 |

| | | | | | | | | | |
|---|---|---|---|---|---|---|---|---|---|
| 88118660 | 16 | c.1606C>T | p.Gln536Ter | LP | JBTS | Dandy-Walker malformation and Retinal Dystrophy | **c.1606C>T, p.Gln646Ter** | **c.1606C>T, p.Gln646Ter** | https://www.ncbi.nlm.nih.gov/pmc/articles/PMC4643834/ |
| 88118673 | 16 | c.1593C>A | p.Tyr531Ter | P | not provided | LCA | c.1593C>A, p.Tyr531Ter | c.2T>A, p.Met1Lys | https://onlinelibrary.wiley.com/doi/epdf/10.1002/humu.9485 |
| 88118715 | 16 | c.1550del | p.Asp516_Leu517insTer | not provided | not provided | LCA | c.1550del, p.Leu517Ter | c.2991+1665A>G, p.Cys998Ter | https://www.cell.com/ajhg/fulltext/S0002-9297(07)62755-4 |
| 88120162 | 15 | c.1474A>T | p.Lys492Ter | P | NPHP, MKS, JBTS | N/A | | | |
| 88120207 | 15 | c.1429C>T | p.Arg447Ter | P/LP | SLS, JBTS, MKS, LCA, BBS, NPHP | LCA | c.1429C>T, p.Arg447Ter | c.2991+1665A>G, p.Cys998Ter | https://www.ncbi.nlm.nih.gov/pmc/articles/PMC3625363/ |
| 88120243 | 15 | c.1393del | p.Glu464_Ile465insTer | P | NPHP, JBTS, MKS | N/A | | | |
| 88120246 | 15 | c.1390G>T | p.Glu464Ter | P | JBTS, MKS, NPHP | N/A | | | |
| 88125357 | 13 | c.1078C>T | p.Arg360Ter | P | NPHP, MKS, JBTS, LCA, SLS, BBS | LCA | c.1078C>T, p.Arg360Ter | c.5587-1G>C | https://www.ncbi.nlm.nih.gov/pmc/articles/PMC3283211/ |
| | | | | | | RP | c.1078C>T, p.Arg360Ter | c.6851_6855del, p.Thr228AsnfsTer | https://iovs.arvojournals.org/article.aspx?articleid=2166376 |
| 88125363 | 13 | c.1072C>T | p.Gln358Ter | P | JBTS, NPHP, MKS | N/A | | | |
| 88129007 | 11 | c.881C>G | p.Ser294Ter | P | JBTS | N/A | | | |
| 88129717 | 10 | c.829G>T | p.Glu277Ter | LP | LCA | N/A | | | |
| 88130283 | 9 | c.654T>G | p.Tyr218Ter | P | MKS, JBTS | JSRD | c.654T>G, p.Tyr218Ter | c.4723A>T, p.Lys1575Ter | https://pubmed.ncbi.nlm.nih.gov/26092869/ |
| | | | | | | JSRD | c.654T>G, p.Tyr218Ter | c.5668G>T, p.Gly1890Ter | https://pubmed.ncbi.nlm.nih.gov/26092869/ |
| 88130309 | 9 | c.628A>T | p.Lys210Ter | LP | not provided | N/A | | | |
| 88130324 | 9 | c.613C>T | p.Arg205Ter | P | MKS, JBTS, NPHP | MKS | **c.613C>T, p.Arg205Ter** | **c.613C>T, p.Arg205Ter** | https://www.ncbi.nlm.nih.gov/pmc/articles/PMC2718326/ |
| 88130553 | 8 | c.508A>T | p.Lys170Ter | P | JBTS, NPHP, MKS, Retinal Dystrophy | SECORD | c.508A>T, p.Lys170Ter | c.5182G>T, p.Glu1728Ter | https://www.ncbi.nlm.nih.gov/pmc/articles/PMC5565704/ |
| | | | | | | LCA | c.508A>T, p.Lys170Ter | c.4090G>T, p.Glu1364Ter | https://pubmed.ncbi.nlm.nih.gov/29771326/ |
| 88131209 | 7 | c.451C>T | p.Arg151Ter | P/LP | NPHP, MKS, JBTS | LCA | c.451C>T, p.Arg151Ter | c.2991+1665A>G, p.Cys998Ter | https://iovs.arvojournals.org/article.aspx?articleid=2126833 |
| | | | | | | Retinal Dystrophy | c.451C>T, p.Arg151Ter | c.4723A>T, p.Lys1575Ter | https://pubmed.ncbi.nlm.nih.gov/28829391/ |

| | | | | | | | | | |
|---|---|---|---|---|---|---|---|---|---|
| | | | | | | LCA | c.451C>T, p.Arg151Ter | c.3181_3182delAT, p.Met1061fs | https://jmg.bmj.com/content/53/11/761.long |
| 88136717 | 6 | c.367C>T | p.Gln123Ter | P | JBTS, MKS, NPHP | LCA | c.367C>T, p.Gln123Ter | ? | https://pubmed.ncbi.nlm.nih.gov/21602930/ |
| | | | | | | LCA | c..367C>T, p.Gln123Ter | c.1627G>T, p.Glu543Ter | https://iovs.arvojournals.org/article.aspx?articleid=2624456 |
| 88136762 | 6 | c.322C>T | p.Arg108Ter | P | JBTS, NPHP, MKS, Nyctalopia | LCA | c.322C>T, p.Arg108Ter | c.2991+1665A>G, p.Cys998Ter | https://www.ncbi.nlm.nih.gov/pmc/articles/PMC3048164/ |
| 88139153 | 5 | c.289G>T | p.Glu97Ter | P | MKS, JBTS, NPHP, MKS | MKS | c.289G>T, p.Glu97Ter | c.1984C>T, p.Gln662Ter | https://www.ncbi.nlm.nih.gov/pmc/articles/PMC2718326/ |
| | | | | | | Cone Dystrophy | c.289G>T, p.Glu97Ter | c.5237G>A, p.Arg1746Gln | https://www.ajo.com/article/S0002-9394(19)30551-3/fulltext |
| 88139174 | 5 | c.268A>T | p.Lys90Ter | LP | not provided | N/A | | | |
| 88140970 | 3 | c.166C>T | p.Gln56Ter | P | NPHP, MKS, JBTS | N/A | | | |

Table 1. CEP290 ClinVar listed nonsense variants with published genotypes and associated diseases. Class: P – Pathogenic, LP – Likely Pathogenic, VUS – Variant of Uncertain Significance. Diseases: LCA – Leber's congenital amaurosis, RP – Retinitis Pigmentosa, SECORD – Severe early childhood onset retinal dystrophy, SLS – Senior Loken syndrome, BBS – Bardet-Biedl syndrome, NPHP – Nephronophthisis, JBTS – Joubert syndrome, JSRD – Joubert syndrome and related diseases, MKS – Meckel Gruber syndrome. **Bold font** – homozygous genotypes, *Grey highlight* – Significantly differentially expressed exons. Where genotypes have been published, sources are provided.

**Supplementary Table 2.**

| Variant | Exon Number | HOT-SKIP | Human Splicing Finder | | Splice AI | | | |
|---|---|---|---|---|---|---|---|---|
| | | Δ ESE/ESS | Δ ESE/ESS | Donor or Acceptor sites | Splice acceptor (SA) gain | Splice acceptor (SA) loss | Splice donor (SD) gain | Splice donor (SD) loss |
| c.7153del | 53 | / | **-2** | **Activation of cryptic acceptor site** | 0.00 | 0.00 | 0.00 | 0.00 |
| c.7073_7077dup | 52 | / | **-11** | **Activation of cryptic acceptor site** | 0.00 | 0.01 | 0.00 | 0.00 |
| c.7048C>T | 52 | 0 | **-6** | ns | 0.00 | 0.11 | 0.00 | 0.00 |
| c.6939C>A | 50 | 0 | **-4** | ns | 0.00 | 0.00 | 0.00 | 0.00 |
| c.6892C>T | 50 | 0.2 | **-5** | ns | 0.00 | 0.00 | 0.00 | 0.00 |
| c.6871C>T | 50 | 0.25 | ns | ns | 0.02 | 0.00 | 0.00 | 0.00 |
| c.6836T>A | 50 | -0.54 | ns | **Activation of cryptic donor site** | 0.00 | 0.00 | 0.00 | 0.00 |
| c.6798G>A | 49 | 0 | ns | **ns** | 0.00 | 0.00 | 0.00 | 0.06 |
| c.6797G>A | 49 | 0 | **-4** | **Activation of cryptic acceptor site** | 0.00 | 0.00 | 0.00 | **0.27** |
| c.6634G>T | 48 | **3** | **-8** | ns | 0.00 | 0.00 | 0.03 | 0.04 |
| c.6364A>T | 47 | 0.8 | **-5** | ns | 0.03 | 0.18 | 0.00 | 0.00 |
| c.6331C>T | 46 | 0.67 | **-9** | ns | 0.00 | 0.00 | 0.00 | 0.00 |
| c.6072C>A | 44 | 0 | **-2** | ns | 0.00 | 0.00 | 0.00 | 0.00 |
| c.6031C>T | 44 | 0.14 | **-9** | **Activation of cryptic acceptor site** | 0.00 | 0.01 | 0.00 | 0.00 |
| c.5932C>T | 43 | 0.33 | **-2** | ns | 0.00 | 0.01 | 0.00 | 0.00 |
| c.5803G>T | 42 | -1.08 | **-4** | **Activation of cryptic donor site** | 0.00 | 0.00 | **1.00** | 0.00 |
| c.5788A>T | 42 | **3** | **-9** | ns | 0.00 | 0.00 | 0.00 | 0.00 |
| c.5776C>T | 42 | 0.2 | **-2** | ns | 0.00 | 0.00 | 0.01 | 0.00 |
| c.5745dup | 42 | / | ns | ns | 0.00 | 0.02 | 0.00 | 0.01 |
| c.5707A>T | 41 | 0.4 | **-3** | ns | 0.00 | 0.00 | 0.00 | 0.02 |
| c.5704G>T | 41 | **2** | **-11** | ns | 0.00 | 0.00 | 0.00 | 0.00 |
| c.5668G>T | 41 | -0.13 | ns | **Activation of cryptic donor site** | 0.00 | 0.00 | 0.00 | 0.00 |
| c.5344C>T | 39 | 0.6 | **-6** | ns | 0.00 | 0.00 | 0.02 | 0.00 |

| | | | | | | | | |
|---|---|---|---|---|---|---|---|---|
| c.5212G>T | 38 | 0.14 | **-12** | **Activation of cryptic donor site** | 0.00 | 0.00 | 0.00 | 0.00 |
| c.5182G>T | 38 | 1.14 | **-5** | ns | 0.00 | 0.00 | 0.00 | 0.00 |
| c.4960C>T | 37 | 0 | **-11** | ns | 0.00 | 0.00 | 0.00 | 0.05 |
| c.4882C>T | 37 | 0 | **-11** | ns | 0.00 | 0.01 | 0.00 | 0.00 |
| c.4811G>A | 36 | 0 | ns | ns | 0.00 | 0.00 | 0.00 | 0.00 |
| c.4801C>T | 36 | **-5** | **-4** | ns | 0.00 | 0.00 | 0.00 | 0.01 |
| c.4771C>T | 36 | **-5** | **-4** | ns | 0.00 | 0.00 | 0.00 | 0.07 |
| c.4732G>T | 36 | 1 | **-10** | ns | 0.01 | 0.02 | 0.00 | 0.00 |
| c.4723A>T | 36 | **-13** | **-14** | ns | 0.01 | 0.17 | 0.00 | 0.00 |
| c.4522C>T | 35 | **3.17** | **-7** | ns | 0.19 | 0.00 | 0.00 | 0.00 |
| c.4490C>A | 35 | 0.33 | **-3** | **Activation of cryptic donor site** | 0.00 | 0.00 | 0.02 | 0.00 |
| c.4393C>T | 34 | **2** | **-2** | **Activation of cryptic acceptor site** | 0.00 | 0.00 | 0.00 | 0.02 |
| c.4276_4277del | 33 | / | **-6** | **Activation of cryptic donor and acceptor site** | 0.00 | 0.00 | 0.00 | 0.00 |
| c.4243G>T | 33 | 0.33 | **-11** | **Activation of cryptic donor site** | 0.00 | 0.00 | 0.00 | 0.00 |
| c.4186C>T | 32 | 0 | **-11** | ns | 0.00 | 0.00 | 0.00 | 0.05 |
| c.4040G>A | 32 | 0 | ns | **Activation of cryptic acceptor site** | 0.04 | 0.05 | 0.00 | 0.00 |
| c.3943G>T | 31 | **2.33** | **-13** | **Activation of cryptic donor site** | 0.00 | 0.00 | 0.00 | 0.00 |
| c.3904C>T | 31 | **2.67** | **-9** | **Activation of cryptic donor site** | 0.00 | 0.00 | 0.09 | 0.00 |
| c.3894dup | 31 | / | **-2** | **Activation of cryptic acceptor site** | 0.00 | 0.00 | 0.00 | 0.00 |
| c.3814C>T | 31 | 0 | **-6** | ns | 0.00 | 0.00 | 0.00 | 0.00 |
| c.3802C>T | 31 | **8.67** | **-5** | ns | 0.00 | 0.00 | 0.00 | 0.00 |
| c.3593C>A | 31 | -1 | ns | **Activation of cryptic donor and acceptor site** | 0.01 | 0.00 | 0.00 | 0.00 |
| c.3420T>G | 28 | **-2.92** | **-5** | **Activation of cryptic donor and acceptor site** | | | | |
| c.3220G>T | 28 | -0.14 | **-12** | ns | 0.00 | 0.07 | 0.00 | 0.01 |
| c.3205_3206delinsTG | 28 | / | **-6** | ns | 0.00 | 0.00 | 0.00 | 0.00 |

| | | | | | | | | |
|---|---|---|---|---|---|---|---|---|
| c.3190del | 28 | / | **-2** | ns | 0.02 | 0.00 | 0.00 | 0.00 |
| c.3175del | 28 | / | ns | ns | 0.06 | 0.00 | 0.00 | 0.00 |
| c.3123dup | 28 | / | ns | **Activation of cryptic acceptor site** | | | | |
| c.3097A>T | 27 | -0.19 | **-2** | ns | 0.00 | 0.00 | 0.00 | 0.00 |
| c.2941C>T | 26 | **9.75** | **-4** | **Activation of cryptic donor site** | 0.00 | 0.00 | 0.00 | 0.01 |
| c.2911G>T | 26 | 1 | **-6** | ns | 0.00 | **0.23** | 0.00 | 0.00 |
| c.2722C>T | 25 | 0.22 | ns | **Activation of cryptic donor site** | 0.00 | 0.00 | 0.00 | 0.00 |
| c.2708T>G | 25 | **-3.37** | ns | ns | 0.00 | 0.01 | 0.00 | 0.00 |
| c.2605C>T | 25 | **10** | ns | ns | 0.00 | **0.42** | 0.00 | 0.00 |
| c.2351T>A | 22 | **-4** | ns | ns | 0.00 | 0.00 | 0.03 | 0.04 |
| c.2251C>T | 22 | -1.25 | ns | ns | 0.00 | **0.27** | 0.00 | 0.00 |
| c.2249T>G | 22 | **-5.5** | **3** | ns | 0.11 | 0.00 | 0.00 | 0.00 |
| c.1984C>T | 20 | 0 | ns | **Activation of cryptic donor site** | 0.00 | 0.00 | 0.00 | 0.00 |
| c.1936C>T | 20 | 0 | ns | **Activation of cryptic donor site** | 0.00 | **0.27** | 0.00 | 0.01 |
| c.1858G>T | 19 | 0.13 | **-16** | ns | 0.00 | 0.02 | 0.00 | 0.00 |
| c.1781T>A | 18 | -0.11 | ns | ns | 0.00 | 0.01 | 0.00 | **0.63** |
| c.1709C>G | 17 | 0 | ns | ns | 0.00 | 0.00 | 0.01 | 0.00 |
| c.1645C>T | 17 | 0 | ns | ns | 0.01 | 0.00 | 0.00 | 0.00 |
| c.1627G>T | 17 | 0.25 | ns | ns | 0.01 | 0.00 | 0.00 | 0.00 |
| c.1606C>T | 16 | 0 | **-6** | ns | 0.00 | 0.00 | 0.00 | **0.57** |
| c.1593C>A | 16 | 0 | ns | **Activation of cryptic donor site** | 0.00 | 0.00 | 0.00 | **0.25** |
| c.1550del | 16 | / | ns | ns | 0.13 | 0.00 | 0.00 | 0.00 |
| c.1474A>T | 15 | **3.89** | ns | **Activation of cryptic donor and acceptor site** | 0.01 | 0.00 | 0.00 | **0.26** |
| c.1429C>T | 15 | 0 | ns | ns | 0.00 | 0.00 | 0.00 | 0.00 |
| c.1393del | 15 | / | ns | ns | 0.00 | 0.00 | 0.00 | 0.00 |
| c.1390G>T | 15 | 1.25 | **-6** | ns | 0.00 | 0.00 | 0.00 | 0.00 |

| | | | | | | | | |
|---|---|---|---|---|---|---|---|---|
| c.1078C>T | 13 | 0 | ns | ns | 0.03 | 0.00 | 0.00 | 0.00 |
| c.1072C>T | 13 | 0.4 | **-7** | ns | 0.02 | 0.01 | 0.00 | 0.00 |
| c.881C>G | 11 | 0.05 | ns | ns | 0.01 | 0.00 | 0.00 | 0.00 |
| c.829G>T | 10 | **5** | **-16** | ns | 0.00 | 0.00 | 0.10 | 0.02 |
| c.654T>G | 9 | **2** | ns | **Activation of cryptic acceptor site** | 0.00 | 0.00 | 0.00 | 0.02 |
| c.628A>T | 9 | 1 | ns | ns | 0.00 | 0.00 | 0.00 | 0.01 |
| c.613C>T | 9 | 0 | ns | ns | 0.00 | 0.00 | 0.00 | 0.00 |
| c.508A>T | 8 | 0.5 | ns | ns | 0.00 | 0.01 | 0.00 | 0.02 |
| c.451C>T | 7 | 1.2 | ns | ns | 0.00 | **0.33** | 0.00 | **0.28** |
| c.367C>T | 6 | **3.5** | ns | ns | 0.00 | 0.00 | 0.00 | 0.00 |
| c.322C>T | 6 | 0.25 | ns | ns | 0.00 | 0.09 | 0.00 | 0.00 |
| c.289G>T | 5 | 0.18 | ns | ns | 0.01 | 0.07 | 0.00 | 0.13 |
| c.268A>T | 5 | -0.2 | ns | ns | 0.00 | 0.01 | 0.00 | 0.03 |
| c.166C>T | 3 | **6** | **-3** | ns | 0.00 | 0.00 | 0.00 | **0.36** |

Table 2. Splicing predictions for *CEP290* stop variants from three independent splicing software. ESS – exonic splicing silencer, ESE – exonic splicing enhancer. *Highlighted rows*: *grey* – variants in symmetric exons, *dark blue* – variants in exons missing in naturally-occurring protein-producing alternative transcripts, *light blue* – both of the prior highlights apply. *Red font* – 2 or 3 of the *in silico* splicing predictions are significant.
