## Supplementary File 1 for "The role of alternative splicing in CEP290-related disease pathogenesis"

### S2 NCBI SRA Data Information

| Project Accession Number | Tissue Type | Further Notes | Age | Run Accession | Instrument | Library Prep | Paired-end data? | Stranded? | mRNA library | Read Length |
|---|---|---|---|---|---|---|---|---|---|---|
| PRJEB4337 | Kidney | | Adult | ERR315468 | Illumina HiSeq 2000 | Illumina TruSeq v2 | Paired | N | polyA | |
| PRJEB4337 | Kidney | | Adult | ERR315494 | Illumina HiSeq 2000 | Illumina TruSeq v2 | Paired | N | polyA | |
| PRJEB4337 | Kidney | | Adult | ERR315443 | Illumina HiSeq 2000 | Illumina TruSeq v2 | Paired | N | polyA | |
| PRJEB4337 | Kidney | | Adult | ERR315383 | Illumina HiSeq 2000 | Illumina TruSeq v2 | Paired | N | polyA | |
| PRJEB4337 | Brain | | Adult | ERR315477 | Illumina HiSeq 2000 | Illumina TruSeq v2 | Paired | N | polyA | |
| PRJEB4337 | Brain | | Adult | ERR315455 | Illumina HiSeq 2000 | Illumina TruSeq v2 | Paired | N | polyA | |
| PRJEB4337 | Brain | | Adult | ERR315432 | Illumina HiSeq 2000 | Illumina TruSeq v2 | Paired | N | polyA | |
| PRJEB4337 | Thyroid | | Adult | ERR315358 | Illumina HiSeq 2000 | Illumina TruSeq v2 | Paired | N | polyA | |
| PRJEB4337 | Thyroid | | Adult | ERR315412 | Illumina HiSeq 2000 | Illumina TruSeq v2 | Paired | N | polyA | |
| PRJEB4337 | Thyroid | | Adult | ERR315428 | Illumina HiSeq 2000 | Illumina TruSeq v2 | Paired | N | polyA | |
| PRJEB4337 | Thyroid | | Adult | ERR315397 | Illumina HiSeq 2000 | Illumina TruSeq v2 | Paired | N | polyA | |
| PRJEB4337 | Skin | Control | Adult | ERR315401 | Illumina HiSeq 2000 | Illumina TruSeq v2 | Paired | N | polyA | |
| PRJEB4337 | Skin | Control | Adult | ERR315460 | Illumina HiSeq 2000 | Illumina TruSeq v2 | Paired | N | polyA | |
| PRJEB4337 | Skin | Control | Adult | ERR315339 | Illumina HiSeq 2000 | Illumina TruSeq v2 | Paired | N | polyA | |
| PRJNA280600 | Brain | | Adult | SRR1957183 | Illumina HiSeq 2000 | Illumina TruSeq Str | Paired | Y | rRNA depleted | 100bp |
| PRJNA280600 | Cerebellum | | Adult | SRR1957125 | Illumina HiSeq 2000 | Illumina TruSeq Str | Paired | Y | rRNA depleted | 100bp |
| PRJNA393104 | Cerebellum | | Adult | SRR5804447 | Illumina HiSeq 1500 | NEBNext Ultradirec | Paired | Y | Doesn't say | |
| PRJNA393104 | Cerebellum | | Adult | SRR5804438 | Illumina HiSeq 1500 | NEBNext Ultradirec | Paired | Y | Doesn't say | |
| PRJNA393104 | Cerebellum | | Adult | SRR5804423 | Illumina HiSeq 1500 | NEBNext Ultradirec | Paired | Y | Doesn't say | |
| PRJNA476171 | Retina | | Adult | SRR7460846 | Illumina HiSeq 2500 | Illumina TruSeq Str | Paired | Y | rRNA depleted | 125bp |
| PRJNA476171 | Retina | | Adult | SRR7460876 | Illumina HiSeq 2500 | Illumina TruSeq Str | Paired | Y | rRNA depleted | 125bp |
| PRJNA476171 | Retina | | Adult | SRR7460899 | Illumina HiSeq 2501 | Illumina TruSeq Str | Paired | Y | rRNA depleted | 125bp |
| PRJNA476171 | Retina | | Adult | SRR7460957 | Illumina HiSeq 2502 | Illumina TruSeq Str | Paired | Y | rRNA depleted | 125bp |
